# An Audit of Use and Monitoring of ACE-inhibitors in General Practice

**DOI:** 10.1101/2021.05.02.21256485

**Authors:** Fíona Coffey, Colin Bradley, Tadhg-Iarla Curran

## Abstract

**Introduction:** ACE-inhibitors are one of the most widely prescribed drugs in general practice for the treatment of hypertension. They are also one of the most frequent drugs associated with preventable drug-related morbidity incidents and guidelines are in place regarding the monitoring of renal function during treatment with these drugs.

**Aim:** To assess the pattern of prescribing and monitoring of ACE-inhibitors in a general practice setting.

**Methods:** A retrospective medical records review was undertaken in a general practice in the Kerry region. Data collected included the indication for the ACE-inhibitor and whether blood pressure, electrolytes, renal function and glomerular filtration rate were measured within three months after initiation and in the period from January 2010 to June 2011. Data was entered into Microsoft Excel and analysed using SPSS.

**Results:** Data on 285 patients was analysed and showed that 78.2% of patients were on ACE-inhibitors for treatment of blood pressure. Following initiation of the ACE-inhibitor, 94.8% of patients had their blood pressure checked within 3 months, while 37.1% of patients had electrolytes checked, 51% of patients had their renal function checked and 31% of people had their glomerular filtration rate checked in the same time period. In the months from January 2010 to June 2011, 91.2% of patients had their blood pressure checked with 73% having electrolytes checked, 80.4% having their renal function checked and 54.7% had their glomerular filtration rate checked. Of those who had their renal function checked 26.1% had an abnormal creatinine level.

**Conclusion:** Follow up of monitoring blood pressure, renal function, electrolytes and glomerular filtration rate within 3 months initiation of the ACE-inhibitor was poor but annual check up scored highly. Having a recall system in place within the practice for reminding patients to get their blood levels checked may help to increase all percentages to 100%.

## Introduction

A normal blood pressure is defined as a systolic pressure of 120 mmHg and a diastolic pressure of 80 mmHg. High blood pressure or hypertension is a repeatedly elevated blood pressure with a systolic pressure above 140 and a diastolic pressure above 90.^1^. The Irish Heart Foundation estimates in Ireland that 10,000 people die each year from cardiovascular disease. It is the most common cause of death, accounting for 36% of all deaths and 22% of premature deaths (under the age of 65). ^2^ Hypertension is a major risk factor for cardiovascular disease and causes many complications such as myocardial infarction, aneurysms, heart failure and renal failure. According to the Institute of Public Health in Ireland, approximately 25.1% of the adult population suffer from hypertension. In 2007, 852,000 Irish adults were recorded as being hypertensive. This number is expected to increase by 40% by the year 2020 to over 1 million suffers.^3^ Hypertension is a modifiable risk factor for cardiovascular disease but despite advances in availability of effective treatment strategies, there is still inadequate control of blood pressure in the hypertensive population.

The NICE guidelines recommend that drug therapy should be offered to patients with a persistent high blood pressure of equal to or greater than 160/100 mmHg and patients who are at an increased risk of cardiovascular disease and have a persistent blood pressure of more than 140/90 mmHg.^4^ The main drugs prescribed for hypertension are angiotensin converting enzyme inhibitors (ACE-inhibitors), angiotensin II receptor blockers, beta-blockers, diuretics and calcium channel blockers. The General Practice Notebook guidelines^5^ in the UK recommend that one week after initiation of treatment with an ACE-inhibitor, patients should have their blood pressure, renal function and serum potassium levels measured. Following this they should have their renal function and electrolytes measured at least once a year.

ACE-inhibitors are one of the most widely prescribed drugs in general practice for the treatment of hypertension and there is a considerable body of evidence in the literature to guide physicians in their use of this class of drugs. A study in 2007 assessing the primary care prescribing patterns in Ireland after the publication of large hypertension trials ^5^ found that the use of ACE-inhibitors continued to rise even after the publication of the ALLHAT ^6^ study in 2002. The ALLHAT study had indicated that thiazide-type diuretics should be the drug of choice for initial treatment of hypertension in most patients requiring drug therapy. It is thought the influence of earlier trials such as the HOPE trial ^8^ and the EUROPA trial ^9^ reinforced the beneficial effect of ACE-inhibitors and the ALLHAT study findings were not actively promoted in primary care in Ireland. ^6^ The HOPE trial found a beneficial effect of treatment with ramipril, a long-acting angiotensin-converting–enzyme inhibitor, among certain predefined subgroups as compared with patients in the placebo group. The HOPE trail concluded that treating 1000 patients with ramipril for four years prevents approximately 150 events in approximately 70 patients. ^8^ In 2006, NICE guidelines recommended that for patients under the age of 55 an ACE-inhibitor should be used as first line treatment.^10^ In 2009, Zaharan et al published a study examining the level of awareness of blood pressure in the population and also ascertained the opinion of general practitioners in the diagnosis and management of hypertension. ^11^ They found that the first choice antihypertensive agent for uncomplicated hypertension in younger patients (40 years old) were ACE-inhibitors. In older patients (70 years old), the first choice antihypertensive agent was reported as diuretics followed by ACE-inhibitors.

### ACE-Inhibitors and Renal Function

Acute kidney injury is defined as an abrupt reduction in renal function, usually heralded by a rise in serum creatinine concentration. After the introduction of captopril, an angiotensin-converting-enzyme inhibitor, in the late 1970s, a syndrome of acute reversible renal failure was described in anecdotal case reports and in a small series of patients receiving ACE-inhibitors. Usually, renal insufficiency was reversed after withdrawal of the offending drug. This indicated that there was a functional, rather than a nephrotoxic, basis for the decline in glomerular filtration rate. ^12^ This was proven by Hricik et al who demonstrated that renal insufficiency due to ACE-inhibitors is the result of intrarenal haemodynamic disturbances.^13^

In normal kidneys, inhibition of the renin-angiotensin system is followed by a substantial increase in renal blood flow and a decrease in the filtration fraction while the glomerular filtration rate remains unchanged. This is due to a suppression of the vasoconstrictive effect of angiotensin II on the efferent arterioles of the glomeruli.^14^ In sodium depleted patients, renal hypoperfusion develops which causes angiotensin II inhibition to lead to a rapid decrease of efferent arteriolar resistances. As a result glomerular filtration rate may become deeply depressed. ^15^ A study by Bridoux et al in 1991 observed acute renal failure in 27 patients who were treated by various angiotensin-converting-enzyme inhibitors for hypertension, heart failure, or a combination of both. None of the patients had significant renal artery stenosis on angiography. Hypotension was present in 12 of the patients and overt volume depletion in 21 cases. Their results showed that renal insufficiency may occur in a significant number of patients during treatment with ACE-inhibitors without stenosis of the main renal arteries. ^16^ A study by Devoy et al observed 15 patients who presented with deterioration in renal function coincident with the introduction of angiotensin-converting-enyzme inhibitors. Four patients remained dialysis dependent after the drug was removed and died within four weeks of presentation. Five patients required short-term dialysis and the serum creatinine level remained above pre-treatment values in seven patients. They concluded that deterioration in renal function associated with angiotensin-converting-enzyme-inhibitor therapy is not always reversible and ACE-inhibitors should be used with great care in patients in whom atherosclerotic vascular disease is likely. ^17^ This syndrome of “functional renal insufficiency” usually develops shortly after initiation of ACE inhibitor therapy but can be observed after months or years of therapy, even in the absence of prior ill effects.^18^ This is one of the reasons general practitioners must continue to assess renal function in patients on ACE-inhibitor therapy, with or without chronic renal disease.

### ACE-Inhibitors and Preventable Drug-Related Morbidity

The World Health Organisation has defined an adverse event as *“an injury related to medical management, in contrast to complications of disease. Medical management includes all aspects of care, including diagnosis and treatment, failure to diagnose or treat, and the systems and equipment used to deliver care*”. The Clinical Negligence and Access to Justice Conference in April 2010 was told that, based on extrapolating international figures to Ireland, there could be 160,000 adverse events in Irish hospitals per year, causing up to 8,000 deaths. It is estimated that there are around 850 000 adverse events in UK hospitals each year, costing the NHS approximately £100 million in increased hospital stays.^19^ Epidemiological studies have found that the classes of drugs most commonly associated with adverse drug reactions in the elderly include diuretics, warfarin, non-steroidal anti-inflammatory drugs (NSAIDs), selective serotonin reuptake inhibitors, beta-blockers and angiotensin-converting-enzyme (ACE)-inhibitors. ^19^ A study by Morris et al in 2003 examined preventable drug related morbidity events in 8 practices in the North West and East Midlands of England. ^20^ It found that out of a total of 507 events, 84 were caused by raised serum creatinine (>150 mmol/l), especially in patients where the creatinine level was not monitored before starting an ACE-inhibitor, within 6 weeks of commencement and at least annually thereafter. There was also 61 events of hyperkalaemia (potassium level >5.5mmol/l) which was caused by use of an ACE-inhibitor for the same reasons as above. In both cases, all eight practices had at least one event. It was noted a small number of drugs contributed to approximately 60% of the events. This included the prescribing of NSAIDS, hypnotic-anxiolytics and ACE-inhibitors.

Currently there is little evidence of the pattern of monitoring of ACE-inhibitors in Ireland. It is hoped that this study may prompt the development of national guidelines for the use and monitoring of ACE-inhibitors.

## AIM

The primary aim of this audit was to evaluate the prescribing and monitoring of ACE-inhibitors in a general practice setting.

The secondary aim was to compare different demographics, such as age, sex, status of the patient and the presence or absence of diabetes mellitus, on the prescribing and monitoring of ACE-inhibitors.

Objectives

The aims were met by carrying out the following objectives:

- Obtaining project and ethical approval to be allowed to carry out the audit
- Obtaining permission from the partners of the general practice to be allowed to use their patients’ files in this current study.
- Identifying patients of the practice who were currently prescribed ACE-inhibitors
- Constructing a database to securely record all data obtained
- Reviewing the patient’s file to obtain data regarding when the ACE-inhibitor was initiated, the indication for initiation and which ACE-inhibitor was selected
- Recording whether blood pressure, electrolytes, renal function and glomerular filtration rate were measured in the three months following the initiation of the ACE-inhibitor
- Recording whether blood pressure, electrolytes, renal function and glomerular filtration rate were measure in the period from the 1^st^ January 2010 to the 1^st^ of June 2011
- Recording the latest blood pressure for the patient
- Recording whether the patient was diabetic or not
- Performing statistical analysis of the data obtained

## Methods

The first step was an initial discussion regarding the project questionnaire with the Professor of General Practice to decide the most appropriate and relevant questions for this audit. The general practice in which the audit was to be performed was visited in order to discuss the proposal and any queries or worries that they may have. Once permission was granted by the general practice to use their files, both project and ethical approval were applied for.

## Ethical Approval

The proposed audit was considered by the Clinical Research Ethics Committee (CREC). Ethical approval was provisionally granted on 13^th^ July 2011 (Appendix One). After applying my sample questionnaire to a selection of medical files, the final questionnaire was decided upon and full ethical approval was applied for which was granted on 1^st^ November 2011 (Appendix Two).

## STUDY DESIGN

### Study Population

The audit was set in a busy 6-doctor general practice in Kerry. As the audit involved the retrospective study of medical files there was no need to recruit any participants once approval and permission of the general practice had been granted. The following inclusion and exclusion criteria were applied to all files in the practice.

### Inclusion criteria

- All patients who are currently receiving a prescription for an ACE-inhibitor from any doctor at the practice
- The patient must still be attending the practice in June 2011
- Any patient who may have been prescribed an ACE-inhibitor prior to joining this practice but were still receiving the ACE-inhibitor from a doctor of this practice up to June 2011

### Exclusion criteria

- Any patient who was no longer a patient of the practice due to a change of location, a change of GP or death of the patient
- Any patient who resided in a nursing home or community hospital
- Any patient who attended a cardiac clinic for follow up

HEALTH *one* was used to search for patients who were currently prescribed an ACE-inhibitor. The British National Formulary (BNF) and Medial Information Management

System (MIMMS) were used in order to search for all ACE-inhibitors currently available in Ireland.

### Study Measures

The questionnaire was designed following NICE guidelines for the prescribing of ACE-inhibitors and the data was gathered by filling the questionnaire (Appendix Three). An initial pilot study of 10 patients was carried out prior to the main study. Following this, the questionnaire was reviewed and modified to allow for further relevant data to be collected.

### Procedures

Each drug was searched separately on HEALTH *one* which provided a list of patient names, date of birth and MRN number. This was repeated 41 times for each ACE-inhibitor available in Ireland as drugs were recorded by brand name rather than generic name. The practice only prescribed 13 different trade name drugs of which there were seven different generic ACE-inhibitors. After applying my inclusion and exclusion criteria, there were 285 remaining patients.

## AGE/SEX/PATIENT STATUS

Data regarding the patient’s age and gender was collected from their medical file. It was also evident on the file as to whether the patient was a private patient or a medical card holder.

## ACE-INHIBITOR

The year the ACE-inhibitor was initiated was identified by accessing the patient’s prescription history and recording the date the ACE-inhibitor was first prescribed.

This was double checked by comparing the prescription history to the transaction history for that date. The transaction history also gave the reason why the ACE-inhibitor was initiated. In cases where the first prescribing of the ACE-inhibitor corresponded to the first visit of the patient, the year the ACE-inhibitor was initiated could not be recorded. This also meant that in some cases the reason for initiation of the ACE-inhibitor could not be recorded either, although in a select number of cases, a thorough history taken by the doctor gave details as to why the ACE-inhibitor was begun. There were a few cases where there was no clear indication as to why to the ACE-inhibitor was started and so the indication was recorded as unknown.

## BLOOD PRESSURE

Blood pressure was recorded as being assessed if a systolic and diastolic reading was visible in the transaction history for the specific time periods being measured. The transaction history was reviewed for the three months following the initiation of the ACE-inhibitor on a transaction by transaction basis. The same method was applied to the 18 month period from the 1^st^ January 2010 to the 1^st^ of June 2011, looking for a systolic and diastolic reading and whether they were within normal limits.

## ELECTROLYTES

Data regarding electrolytes was collected by searching the laboratory results in the three month period following the initiation of the ACE-inhibitor and the 18 month period from the 1^st^ January 2010 to the 1^st^ of June 2011. If blood results showing sodium and potassium levels were present then it was accepted that electrolyte levels had been assessed. A blood results showing an abnormal sodium or potassium level (usually highlighted in red), was recorded as not being within normal limits.

## RENAL FUNCTION

Renal function status was measured by looking at the laboratory results in the three month period following the initiation of the ACE-inhibitor and the 18 month period from the 1^st^ January 2010 to the 1^st^ of June 2011. If blood results showing urea and creatinine were present then it was accepted that renal function had been assessed and any abnormal results were recorded.

## GLOMERULAR FILTRATION RATE

Glomerular filtration rate is estimated using an algorithm based on the patient’s age, gender, creatinine level and race. There is a tool on HEALTH *one* to calculate this figure or alternatively it can also be calculated using an online calculator. Glomerular filtration rate monitoring was assessed by reviewing all transactions during the three month period following the initiation of the ACE-inhibitor and the 18 month period from the 1^st^ of January 2010 to the 1^st^ of June 2011 and noting whether glomerular filtration rate or ‘eGFR’ was recorded. As well, the tool on HEALTH *one* for calculating glomerular filtration rate was reviewed as it stores all calculations in chronological order for each patient in their file. If no figure was evident on either of these searches then the glomerular filtration rate was recorded as having not been assessed.

## MOST RECENT BLOOD PRESSURE READING

The patient’s transactions were reviewed in reverse chronological order looking for the most recent blood pressure reading. Both systolic and diastolic readings were recorded. An 18 month time frame was used as the inclusion criterion for most recent blood pressure reading and any readings beyond this time period were not recorded.

## DIABETES MELLITUS

The patient’s diabetes mellitus status was assessed by looking at the patient’s past medical history and medications combined, from the start of each patient’s transaction history. The patient was recorded as either suffering from diabetes mellitus or not suffering from diabetes mellitus. Both diabetes mellitus type one and type two were included.

## LIMITATIONS

In 75 cases, it was not possible to record when the patient started the ACE-inhibitor as computerised files were added to the practice around 1998-1999. In these cases it was also not possible to record whether blood pressure, electrolytes, renal function and glomerular filtration rate were assessed within three months of initiation of the ACE-inhibitor and in many cases the indication for the ACE-inhibitor. Likewise, this occurred with patients who joined the practice after starting on ACE-inhibitor from a doctor outside the practice. This was adjusted in the data analysis.

### Timetable

Please see Appendix Four.

### Security

Patients’ names, date of birth and MRNs were excluded and all files and data were password protected.

## Data Analysis

Data was gathered onto Microsoft Word 2007 questionnaire and transferred over to a database created on Microsoft Excel 2007. All work was carried out on a Packard Bell Easynote TJ65 workstation. The data was then exported to Predictive Analytics Software (PASW) version 18 to analyse any findings. The values of variables were re-coded and re-grouped where necessary. A split file function was used to compare different groups.

Descriptive and inferential statistics, frequencies and graphs were produced.

## RESULTS

Firstly the clinical characteristics and prescribing pattern of the study will be discussed with emphasis on the indication for the ACE-inhibitor and the most popular ACE-inhibitor prescribed. Secondly the results of monitoring after initiation and monitoring in the time period from the 1^st^ of January 2010 to the 1^st^ of June 2011 will be reported on. This will include any abnormal electrolyte and renal function results. Thirdly any differences in monitoring in the time period from the 1^st^ of January 2010 to the 1^st^ of June 2011 between different demographics such as age, patient status and diabetic status will be described. Finally the results of the most recent blood pressure reading will be discussed.

## Clinical Characteristics

285 patients fitted the criteria and were used in the audit. 52.63% of patients receiving an ACE-inhibitors were male. The mean age of patients receiving and ACE-inhibitor was 65.87 years old, with a range from 23 to 95 years of age. With regards to patient status, 69.96% of the patients analysed were medical card holders. 19.57% of patients receiving ACE-inhibitors were diabetic, type I and type II diabetes mellitus were included in this figure.

## Prescribing Pattern

**Graph 1.**
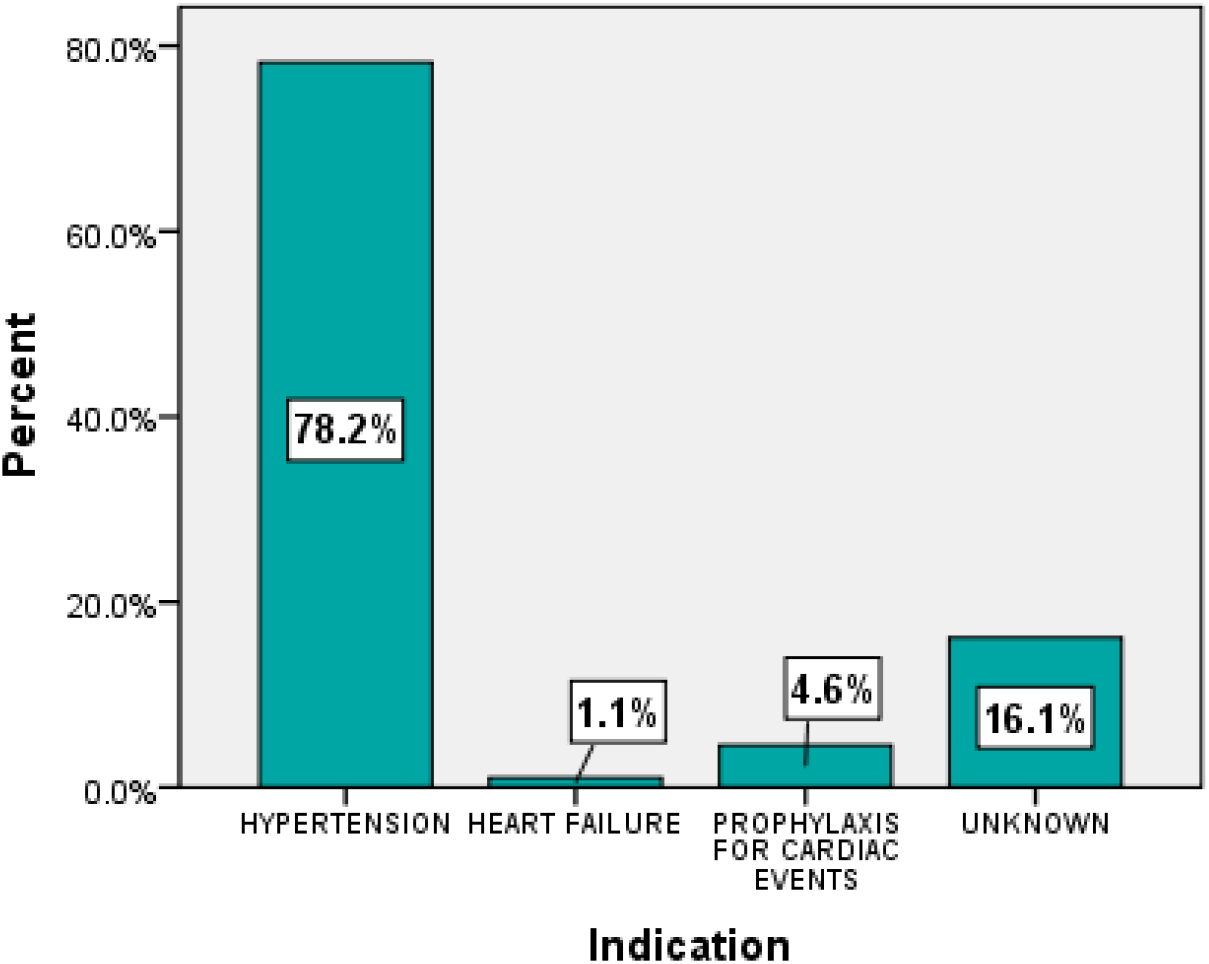
Indication for ACE-inhibitor.

As evident in Graph 1 above, the main indication for the prescribing of ACE-inhibitors was the treatment of hypertension. In 16.1% of cases the reason for the ACE-inhibitor being initiated was unknown and the reasons for this have been discussed in the method section.

There were seven generic ACE-inhibitors prescribed by doctors in the practice and in alphabetical order these were: captopril, cilazaprile, enalapril maleate, lisinopril, perindopril erbumine, quinapril and ramipril. The most popular ACE-inhibitor was ramipril, prescribed for 78.3% of patients on an ACE-inhibitor in either generic or trade name form (Tritace, Ramilo). Lisinopril was the second most popular at 7.4%, which was closely followed by enalapril at 6%. There was no difference between genders or between diabetic patient and non-diabetic patients with regard to the indication for the ACE-inhibitor or the choice of ACE-inhibitor used. Equally, the status of the patient made no difference in the indication for the ACE-inhibitor. Ramipril was still the most commonly prescribed ACE-inhibitor for both medical card patients and private patients, however 57.6% of medical card patients receiving ramipril were prescribed generic ramipril while 52.9% of private patients receiving ramipril were prescribed trade name drugs, either Ramilo or Tritace. An Independant samples T-test gave a p-value of 0.073 indicating this is not statistically significant.

## MONITORING

### Within Three Months Of Initiation

The corrected values in Table 1 are corrected for unavailable data. The reason for data being unavailable has been discussed in the methods section.

**Table 1.** Monitoring within 3 months of initiation of an ACE-inhibitor.

|  | YES<br>(corrected) | NO<br>(corrected) | NOT AVAILABLE<br>(uncorrected) |
| --- | --- | --- | --- |
| *Blood Pressure* | 94.8% | 5.2% | 26.3% |
| *Electrolytes* | 37.6% | 62.4% | 26.3% |
| *Renal Function* | 51% | 49% | 26.3% |
| *eGFR* | 31% | 69% | 26.3% |

As we can see from Table 1, blood pressure monitoring after initiation scored highly with over 90% of patients having their blood pressure checked within three months of initiation of an ACE-inhibitor. Renal function was measured in just over half the patients in the same time period. Electrolytes and glomerular filtration rate measurements scored poorly, with 37.6% of patients having their electrolytes measured and 31% having their glomerular filtration rate measured.

## 1^st^ January 2010 to 1^st^ June 2011

As demonstrated by Table 2 above, the monitoring of blood pressure and renal function in the time period from the 1^st^ of January 2010 to the 1^st^ of June 2011 scored highly. Electrolyte monitoring was acceptable at 73%. Creatinine level, one of the measurements recorded in renal function blood tests, is used in the calculation of glomerular filtration rate. However the glomerular filtration rate again scored poorly compared to the figure present for renal function

**Table 2.** Monitoring from the 1^st^ of January 2010 to 1^st^ of June 2011.

|  | YES | NO | NOT AVAILABLE |
| --- | --- | --- | --- |
| *Blood Pressure* | 91.2% | 7.7% | 1.1% |
| *Electrolytes* | 73% | 25.6% | 1.1% |
| *Renal Function* | 80.7% | 17.9% | 1.1% |
| *eGFR* | 54.7% | 43.9% | 1.1% |

## ABNORMAL RESULTS

**Graph 2.**
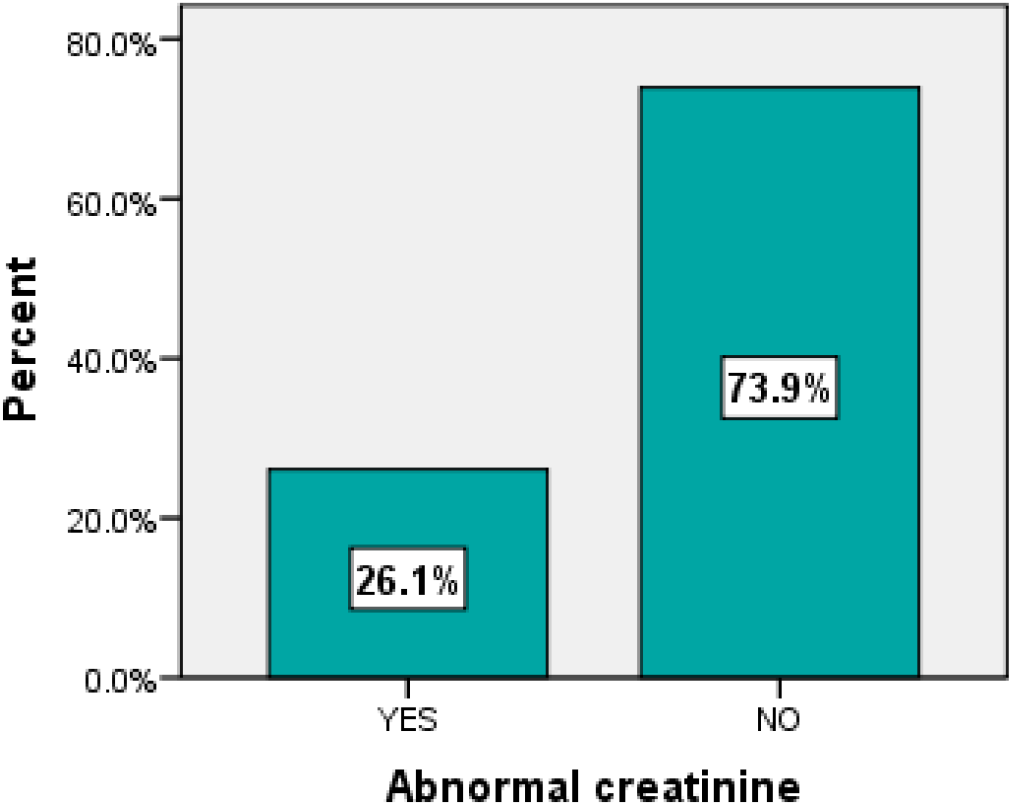
Abnormal creatinine results.

Of the 80.7% of patients who had their renal function checked, 26.1% had an abnormal creatinine level as identified by laboratory results. Of that 26.1% with an abnormal creatinine level, only 68.3% had evidence that their glomerular filtration rate was measured.

208 patients had their electrolytes measured between 1^st^ January 2010 and 1^st^ of June 2011. There were only four abnormal potassium results, hyperkalaemia in all cases.

## DEMOGRAPHICS

### Age

As previously stated the mean age of the population studied was 65.87 years but the range was from 23 to 95 years. In order to assess any differences in monitoring, the population was grouped into different age brackets, up to 40 years, 41 to 50 years, 51 to 60 years, 61 to 70 years, 71 to 80 years, 81 to 90 years and 91 and older years.

As evident from table 3 above, patients aged between 71 and 80 years tended to score the highest across all areas being monitored. Patients less than 40 years old were the least monitored in the practice, with only 16.7% of this group have their glomerular filtration rate estimated. Patients aged between 81 and 90 years old, while scoring highly in the monitoring of renal function, scored very poorly in the calculation of glomerular filtration rate. As we can see from Graph 3 below, this group had the second highest abnormal creatinine levels and so would be expected to have increased monitoring of glomerular filtration rate.

**Table 3.** Monitoring between 1^st^ of January 2010 and 1^st^ of June 2011 dependant on age.

|  | Blood Pressure | Electrolytes | Renal Function | eGFR |
| --- | --- | --- | --- | --- |
| Up to 40 years | 66.7% | 33.3% | 50% | 16.7% |
| 41 to 50 years | 91.2% | 61.8% | 70.6% | 51.9% |
| 51 to 60 years | 93.6% | 73.9% | 82.6% | 54.3% |
| 61 to 70 years | 91.9% | 81.8% | 84.8% | 58.6% |
| 71 to 80 years | 96.2% | 84.9% | 92.5% | 64.2% |
| 81 to 90 years | 91.9% | 56.8% | 73% | 43.2% |
| 91 and older | 83.3% | 66.7% | 66.7% | 66.7% |

**Graph 3.**
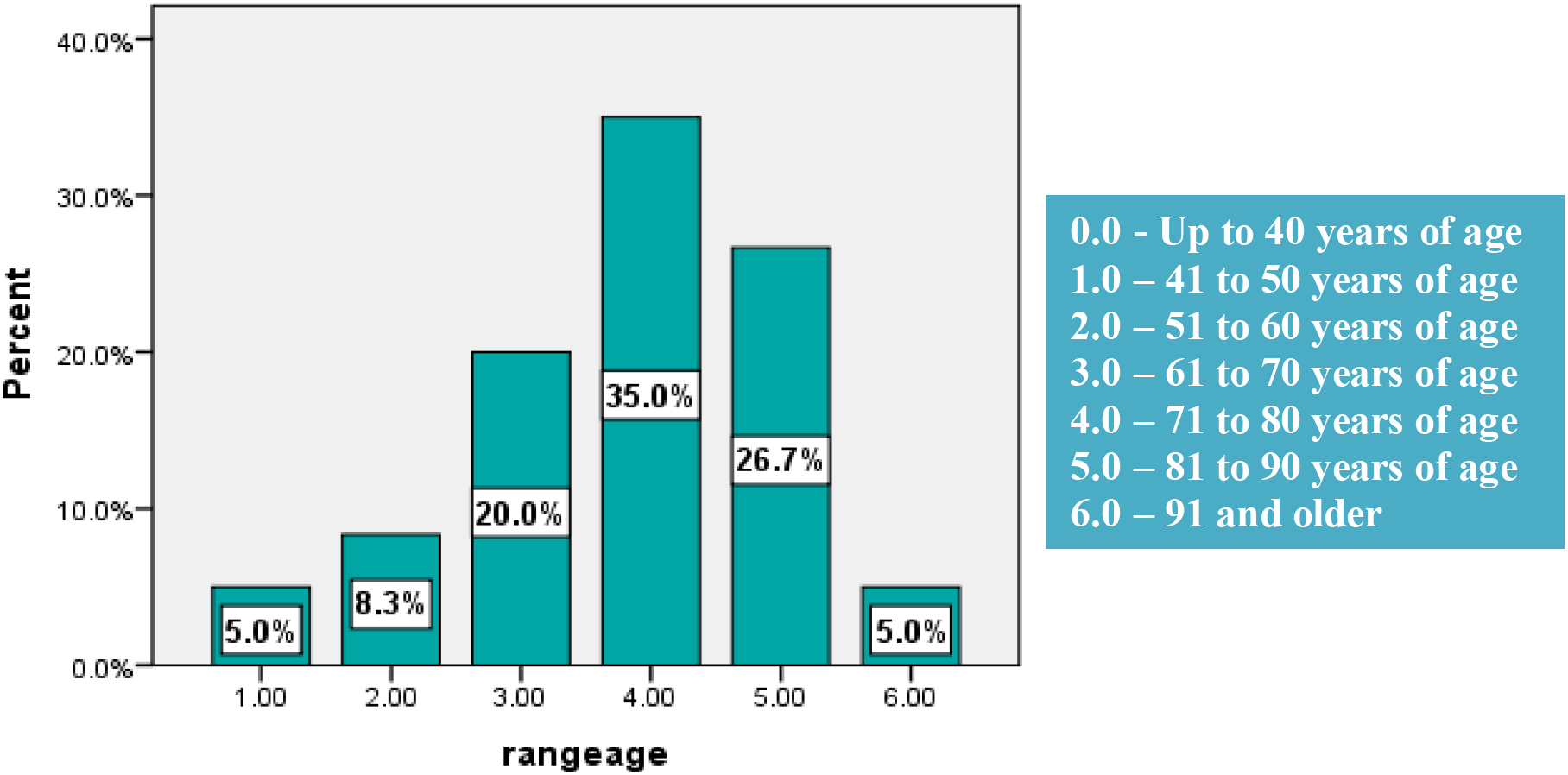
Abnormal creatinine results grouped by age.

As demonstrated by Graph 3 above, group 4.00, patients aged between 71 and 80 years had the highest percentage of abnormal creatinine results. This group had above average monitoring of glomerular filtration rate but at 64.2% it would not be considered ideal.

### Diabetic Patients

Diabetic patients tended to have a higher percentage of monitoring in blood pressure, electrolytes, renal function and glomerular filtration rate. However in the case of blood pressure, electrolytes and renal function, the difference between diabetic patients and non-diabetic patients was minimal, usually around 3%. The most significant difference between diabetic patients and non-diabetic patients in monitoring was with regard to renal function which is demonstrated in Graph 4.

**Graph 4.**
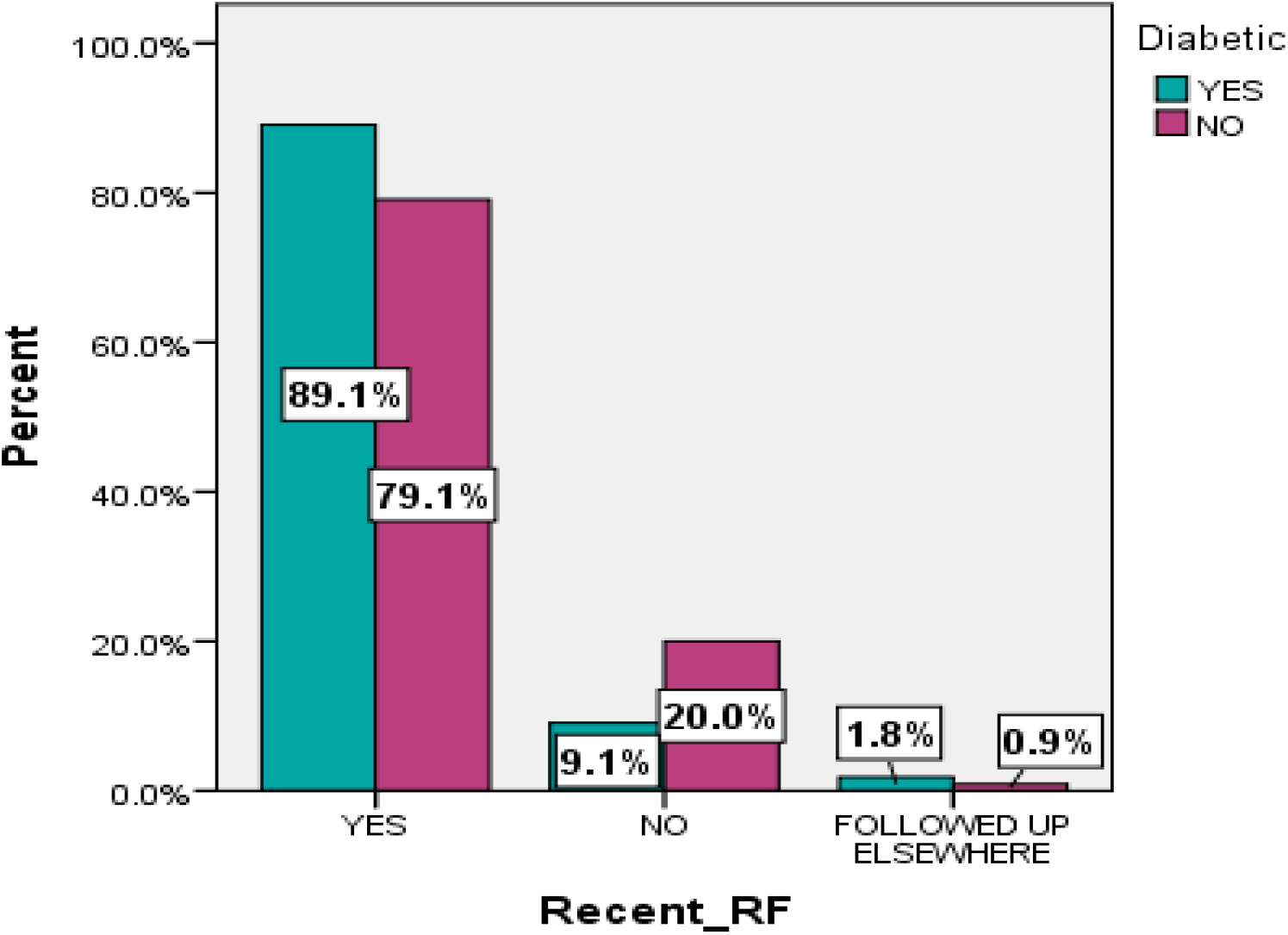
Monitoring of renal function in diabetic patients compared to non-diabetic patients.

As evident by graph 4 above, 89.1% of diabetics have had their renal function checked in the months between 1^st^ of January 2010 to 1^st^ of June 2011 compared with 79.1% of non-diabetics on ACE-inhibitors.

An Independent Sample T test analysing whether there was any significance between diabetics and non-diabetics with regards renal function monitoring in the months between 1^st^ of January 2010 to 1^st^ of June 2011 showed no statistical significance difference.

## Independent Samples Test

|  | Levene's Test<br>forquality of<br>Variances |  | t-test for Equality of Means |  |  |  |  |  |  |
| --- | --- | --- | --- | --- | --- | --- | --- | --- | --- |
|  | F | Sig. | t | df | Sig. (2-tailed) | Mean<br>Differen<br>ce | Std.<br>Error<br>Differen<br>ce | 95% Confidence<br>Interval of the<br>Difference |  |
|  |  |  |  |  |  |  |  | Lower | Upper |
| RF Equal<br>variances<br>assumed | 8.159 | .005 | -<br>1.413 | 278 | .159 | -.091 | .064 | -.217 | .036 |
| Equal<br>variances not<br>assumed |  |  | -<br>1.515 | 90.27<br>1 | .133 | -.091 | .060 | -.209 | .028 |

### Patient Status

As previously stated 69.96% of patients analysed were medical card holders. There was no significant statistical difference in the monitoring of medical card patients and the monitoring of private patients in this audit.

## Most Recent Blood Pressure Reading

The patients most recent blood pressure reading was recorded, both systolic and diastolic levels. As you can see from Graph 5, 45.4% of patients have a systolic blood pressure reading of 131 mmHg or higher and 12.8% had a reading of greater than 150 mm Hg. The diastolic readings, from Graph 6, fared better with 76.9% having a diastolic reading of 80 mmHg or less. This is a once-off reading and does not take into account how much a patient’s blood pressure may have been lowered.

**Graph 5.**
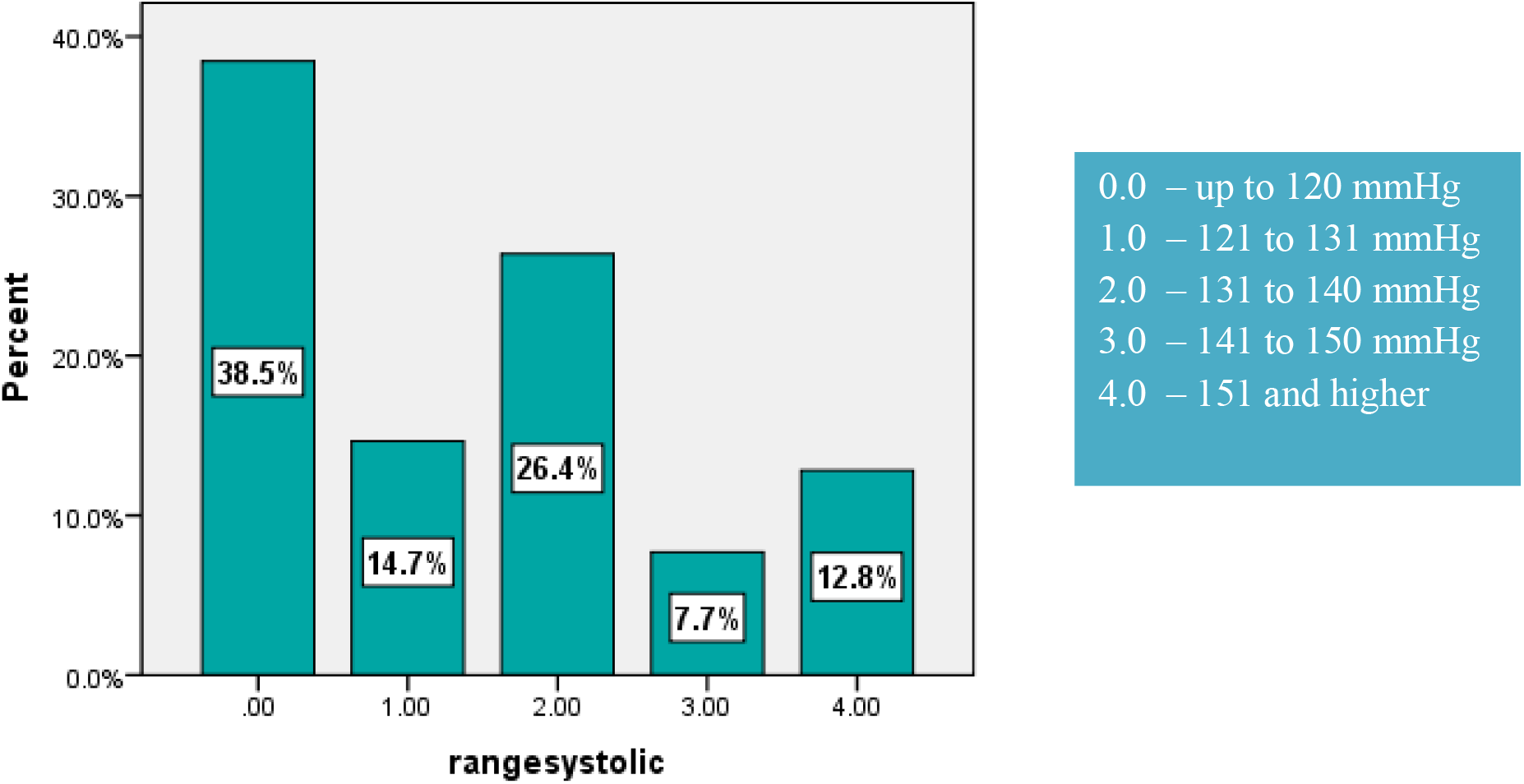
Most recent systolic reading.

**Graph 6.**
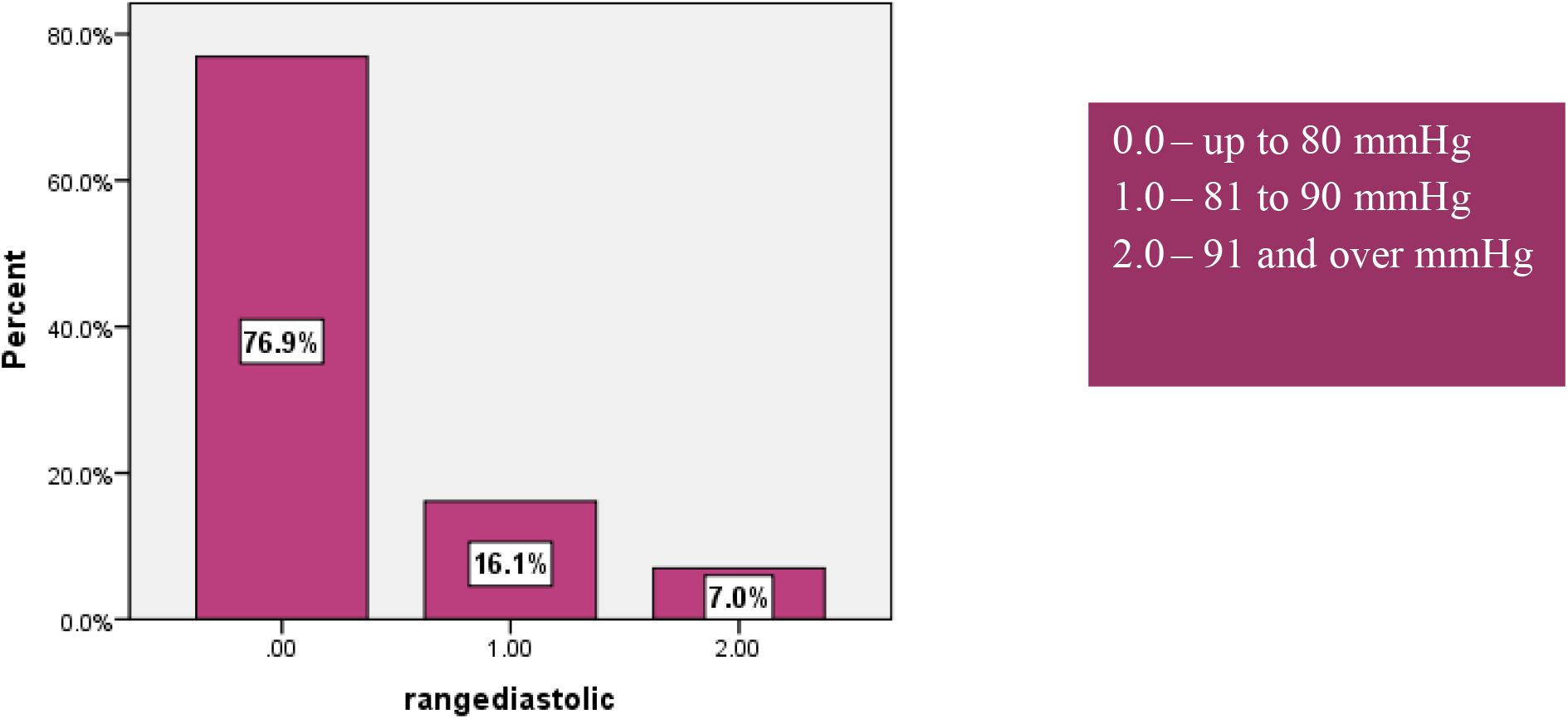
Most recent diastolic reading.

## Discussion

There are many positives to take from this current audit and it also highlights issues with the use and monitoring of ACE-inhibitors that need to be addressed. The Clinical Knowledge Summaries in the UK recommends that ACE-inhibitor selection be limited to the use of lisinopril, ramipril and enalapril.^21^ These were the most popular prescribed ACE-inhibitors in this general practice. There was also no difference in the prescribing or monitoring of private patients compared to medical card patients. Diabetic patients, who are one of the groups that are at an increased risk of renal artery stenosis, were closely monitored with regard to renal function and electrolytes.

With regard to monitoring after initiation of the ACE-inhibitor, there seemed to be no system in place for this and this is one area that needs to be addressed. With an ageing population, and the likelihood of more patients requiring anti-hypertensives ^3^, there must be a clear protocol to follow when prescribing ACE-inhibitors to ensure that all patients are receiving high standards of care. This protocol should be understood and followed by both doctors and nurses and a “recall system” could be introduced into Health *one*.

The monitoring of blood pressure, electrolytes and renal function in the time period between the 1^st^ of January 2010 and the 1^st^ of June 2011 was above average, with blood pressure having been monitored in over 90% of patients. Glomerular Filtration Rate scored poorly compared to renal function. As creatinine level is one of the measurements used in the calculation of glomerular filtration rate one would expect both percentages to match. Most alarming was the fact that 31.7% of patients with an abnormal creatinine level did not have their glomerular filtration rate recorded in their file. Any patient with an abnormal creatinine level should automatically have their glomerular filtration rate calculated and recorded into a transaction in their file. Equally all elderly patients and diabetic patients should have their glomerular filtration rate recorded as they are more at risk of developing acute renal failure.

The audit exposed areas in this practice that need to be looked at and also identified the need for a single set of guidelines for every doctor in the practice to follow.

## Conclusion

ACE-inhibitors are one of the most commonly prescribed drugs for the treatment of hypertension in Ireland. However they have been associated with preventable drug related mortality events. These events usually occur due to insufficient monitoring of serum potassium and creatinine levels. This audit looked at the monitoring of ACE-inhibitors in a busy general practice in Kerry.

The ideal monitoring of ACE-inhibitors has been described as measurement of electrolytes and renal function one week after initiation of an ACE-inhibitor, after every increase in the dosage of the ACE-inhibitor and once a year in patients receiving an ACE-inhibitor.^5^

Monitoring is especially important in patients who are at increased risk of pre-renal acute renal failure, such as patients who suffer from bilateral renal artery stenosis, hypoperfusion, elderly patients and diabetic patients.

In order to ensure that all patients are receiving sufficient monitoring, a protocol drawn up following the most recent guidelines should be adopted by all doctors in the general practice.

Establishing a “recall” system would serve as a reminder to both doctors and nurses to ensure that this monitoring is occurring. A repeat audit after establishment of a “recall” system would identify any further areas that need improvement to ensure that monitoring is occurring in every patient.

### Limitations

Ideally more than one general practice should have been audited to get a clearer idea of the standard of monitoring of ACE-inhibitors and also show how each general practice is performing compared to each other. However, this audit was performed in the biggest general practice in the Killarney region.

The once-off isolated reading of the most recent blood pressure reading would have been strengthened if there was also a recording of blood pressure before initiation to give an idea of the improvement in blood pressure.

A survey of the doctors in the practice could have been performed in order to find their level of knowledge with regard to the guidelines in place for the monitoring of ACE-inhibitors. It would also identify if there was any guideline in particular that they followed which could be incorporated into a single protocol for every doctor in the practice to follow.

## Data Availability

All data referred to in the manuscript was accurate at the time of writing.

## APPENDICES

### Appendix One Provisional Ethical Approval

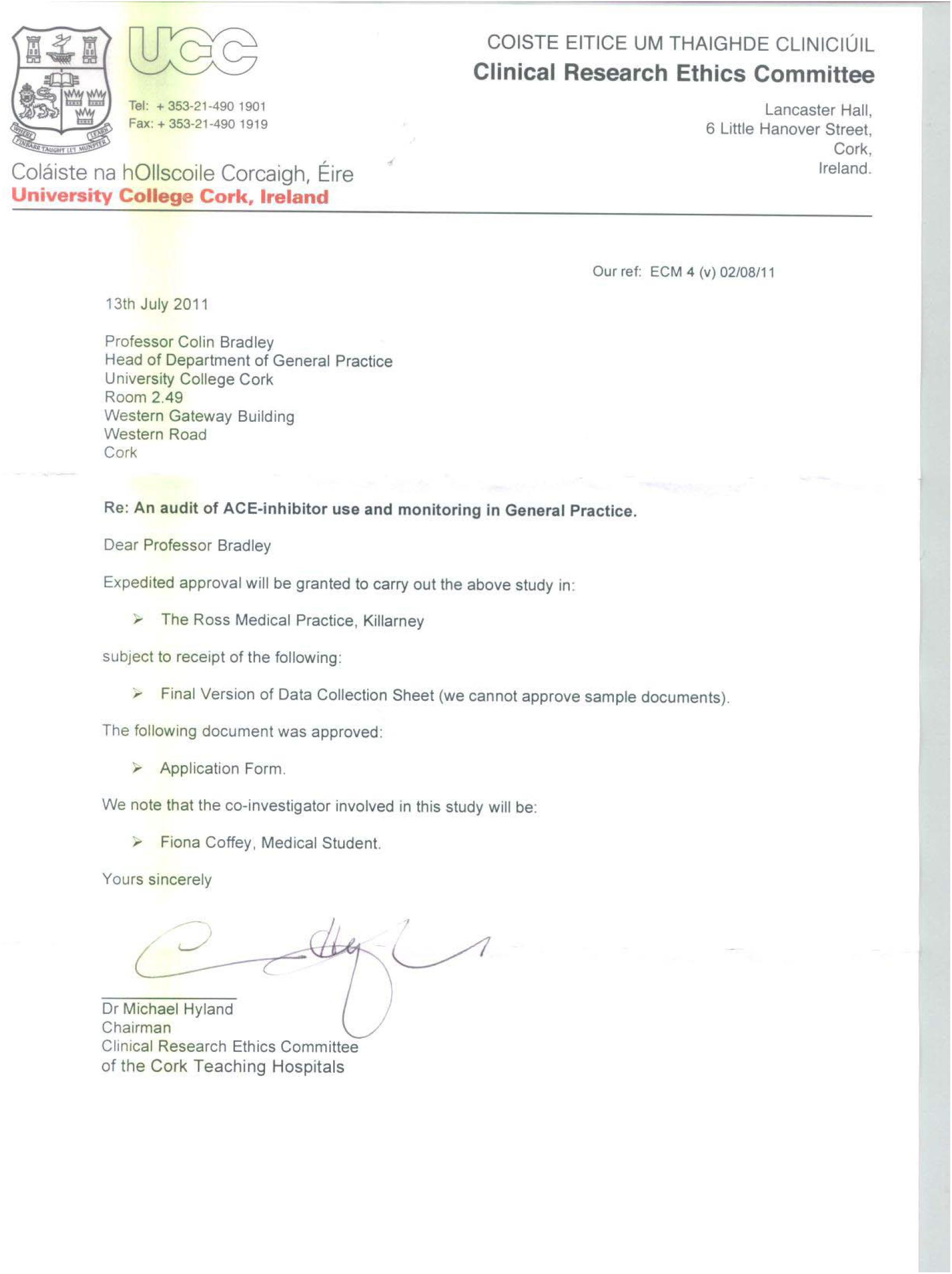

## Appendix Two

### Full Ethical Approval

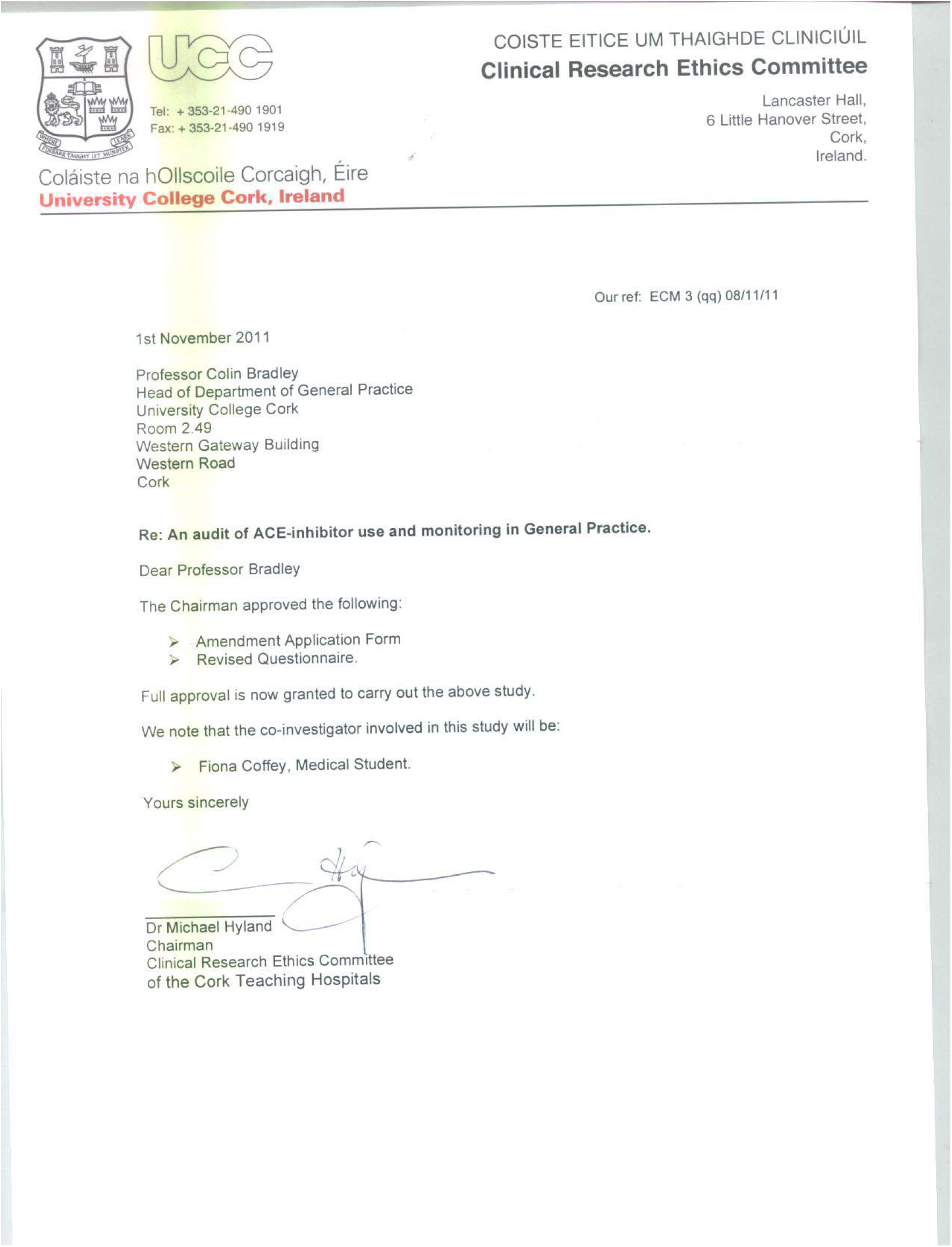

## Appendix Three

### Questionnaire Used for Audit

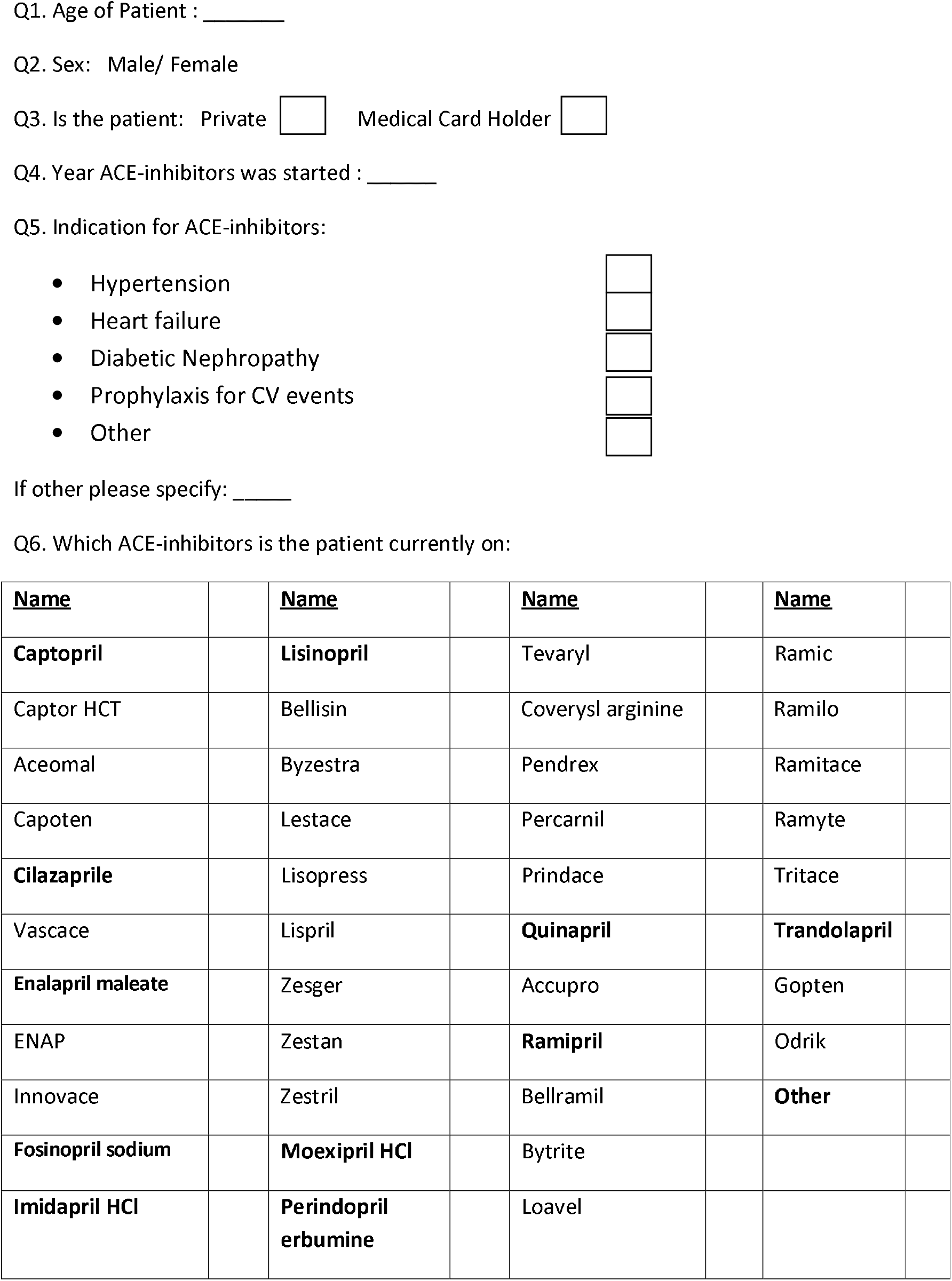
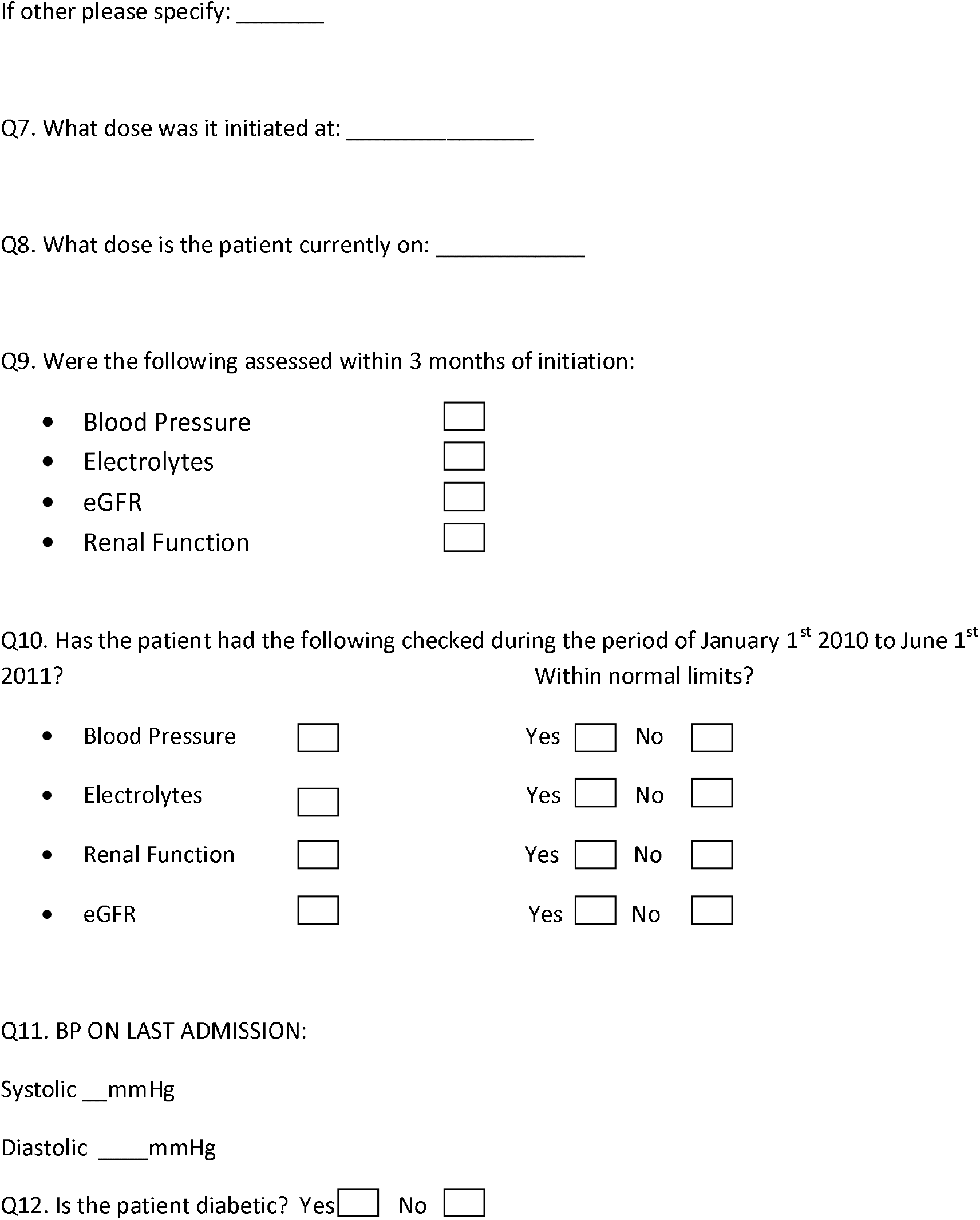

## Appendix Four

### TIMETABLE

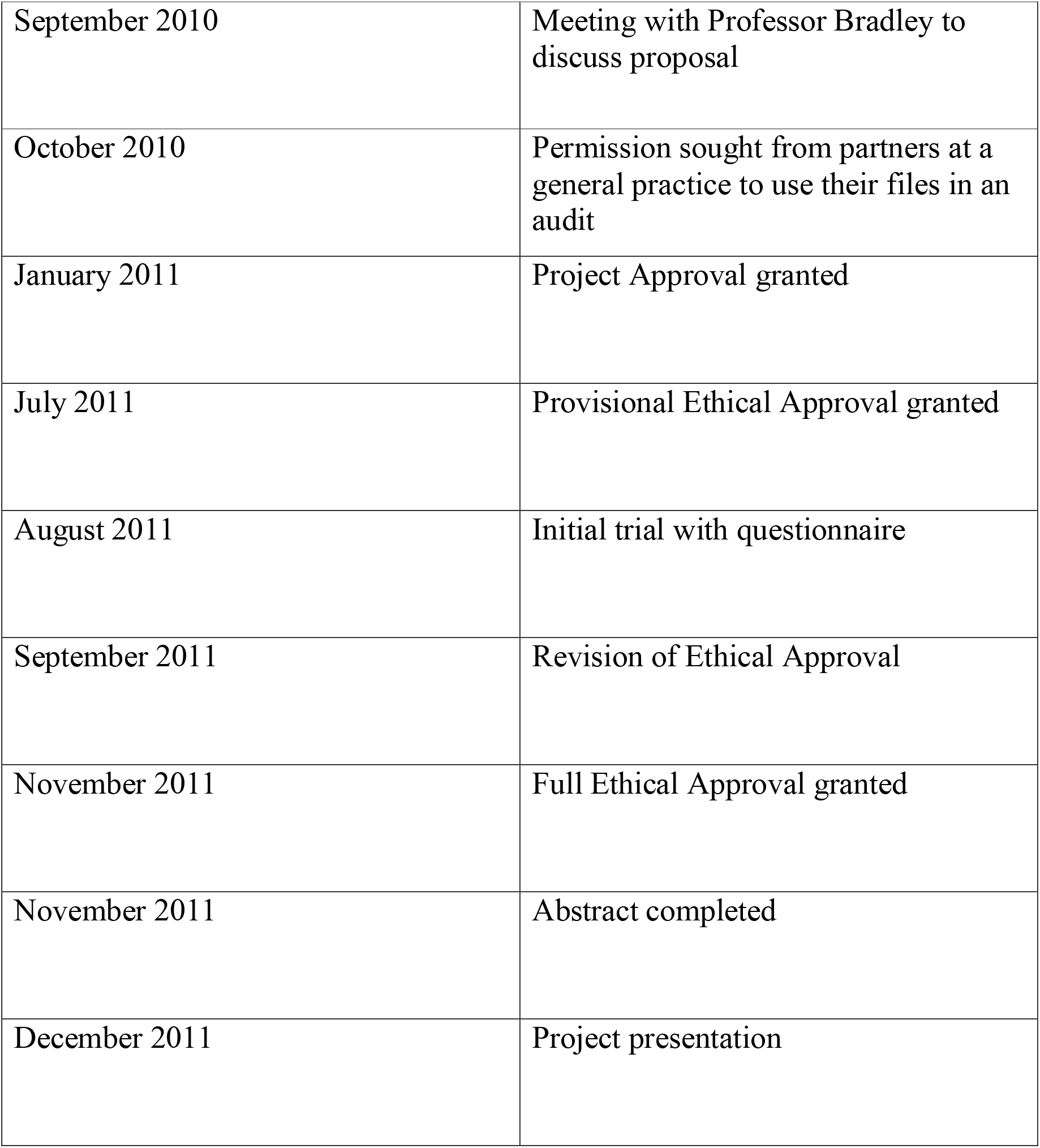

